# Phenotypic overlap between rare disease patients and variant carriers in a large population cohort informs biological mechanisms

**DOI:** 10.1101/2024.04.18.24305861

**Authors:** Lane Fitzsimmons, Undiagnosed Diseases Network, Brett Beaulieu-Jones, Shilpa Nadimpalli Kobren

## Abstract

The biological mechanisms giving rise to the extreme symptoms exhibited by rare disease patients are complex, heterogenous, and difficult to discern. Understanding these mechanisms is critical for developing treatments that address the underlying causes of diseases rather than merely the presenting symptoms. Moreover, the same dysfunctional biological mechanisms implicated in rare recessive diseases may also lead to milder and potentially preventable symptoms in carriers in the general population. Seizures are a common, extreme phenotype that can result from diverse and often elusive biological pathways in patients with ultrarare or undiagnosed disorders. In this pilot study, we present an approach to understand the biological pathways leading to seizures in patients from the Undiagnosed Diseases Network (UDN) by analyzing aggregated genotype and phenotype data from the UK Biobank (UKB). Specifically, we look for enriched phenotypes across UKB participants who harbor rare variants in the same gene known or suspected to be causally implicated in a UDN patient’s recessively manifesting disorder. Analyzing these milder but related associated phenotypes in UKB participants can provide insight into the disease-causing molecular mechanisms at play in the rare disease UDN patient. We present six vignettes of undiagnosed patients experiencing seizures as part of their recessive genetic condition, and we discuss the potential mechanisms underlying the spectrum of symptoms associated with UKB participants to the severe presentations exhibited by UDN patients. We find that in our set of rare disease patients, seizures may result from diverse, multi-step pathways that involve multiple body systems. Analyses of large-scale population cohorts such as the UKB can be a critical tool to further our understanding of rare diseases in general.

## Introduction

Collectively as many as 446 million people, or ~6% of the worldwide population, are afflicted with rare diseases.^1^ However, any specific rare disease may manifest in as few as a handful of patients across the world. Rare diseases are difficult to diagnose, and patients often endure years of arduous, inconclusive testing and misdiagnoses. Even after diagnosis, treatments indicated for these specific diseases are typically limited.^2^ Successful repurposing of other drugs for off-label rare disease treatment is also limited because existing usage guidelines, efficacy, and adverse event measurements from historical clinical studies may not be applicable in rare disease cases.^3^ Despite the critical need for diagnostic and therapeutic improvements for rare disease patients, funders have difficulty justifying resource-intensive research and development when only a small number of patients will benefit. Additionally, support from insurers is often absent or insufficient.^2,4^

Rare diseases are largely genetic in nature. Eighty-five percent are estimated to have genetic etiologies, with the majority classified as recessive conditions.^5^ The manifestation of recessive conditions requires that both copies of relevant autosomal gene(s) or single copies of relevant X-linked gene(s) have been rendered dysfunctional. This can occur when a patient inherits the same deleterious variant from both parents as a homozygous recessive variant.^6^ Sickle-cell anemia, for instance, manifests when both copies of a patient’s *HBB* gene harbor the same A>T base pair mutation at a critical position.^7^ Alternatively, a recessive condition can arise when a patient inherits two different deleterious variants from each parent, called compound heterozygous variants. Cystic fibrosis, for instance, arises when both copies of a patient’s *CFTR* gene are compromised due to potentially different variants inherited from each parent.^8^ In both of these types of recessive cases, affected individuals have seemingly unaffected parents but a potentially significant family history of disease, such as a similarly affected grandparent or sibling.^6^

In addition to known pathogenic variants, other variants in the same disease-causing genes can result in a spectrum of symptoms ranging from benign to pathological. In the *CFTR* gene, for example, nearly 1700 different mutations lead to varying manifestations of cystic fibrosis with differing prognoses and comorbidities.^9^ Intriguingly, recent studies have demonstrated that seemingly healthy individuals in the general population who carry one normally functioning and one compromised copy of the *CFTR* gene may exhibit related yet less severe symptoms typical of cystic fibrosis as well.^10^ Patients with rare genetic disorders often present on the extreme side of the phenotypic spectrum, with debilitating symptoms that severely restrict normal physical and cognitive functioning. The less severe phenotypes exhibited by carriers of variants in the same genes can provide insight into the biological mechanisms at play in recessive disorders. Increased understanding of this disease process would have the potential to both help rare disease patients and potentially lead to preventative treatments in larger “carrier” populations where less severe phenotypes build up over time. For example, if variants in a gene causatively linked to neurodegeneration in a rare disease patient were associated with increased inflammation in the general population, carriers might be at increased risk of developing neurodegenerative conditions as well and could be candidates for clinical studies of early interventions. It can be extremely difficult to gather information and draw conclusions about rare disorders that individually manifest in such small numbers. For suspected genetic disorders where unrelated but similarly-presenting individuals are found, disease-associated genes or genic regions can be identified statistically through case–control analyses. However, for undiagnosed, “N-of-1” cases where no similarly affected individuals are known and no matching disorders have been described in the literature, clinical researchers must rely on external functional information to determine whether and how candidate genes harboring intriguing variants are concordant with a patient’s symptoms.^11,12^ Even once a diagnostic gene is identified, the perturbed mechanisms leading to symptom onset and progression can remain elusive. In these cases, milder disease-related symptoms observed across individuals with only one damaging variant in the same gene may shed light on the underlying biological process or pathway implicated in a rare disease patient’s extreme manifestation of symptoms of the same nature.

The Undiagnosed Diseases Network (UDN) was established in 2014 to serve patients with extremely rare and difficult-to-diagnose conditions. These patients represent “N-of-1” cases, and their disorders or specific phenotypic presentation manifest in so few patients worldwide that it is nearly impossible to first identify similarly presenting patients and then to search for recurrently-implicated, significantly associated genomic loci. The majority of patients who enroll in the UDN suffer from sporadic or suspected recessive conditions that manifest across multiple body systems. Examining these rare and complicated conditions in the context of healthy individuals who carry similar variants has the potential to benefit patients directly by identifying root causes of certain symptoms and to more broadly enhance understandings of biological mechanisms of recessive illnesses. Moreover, establishing connections between rare disorders and potentially actionable symptoms in the general population may further incentivize the development of improved diagnostics and therapeutics for these patients.

Here, we present a framework and pilot study to examine genes likely or known to be implicated in extremely rare disorders through the lens of cohort-level phenotype enrichments. We include in our pilot a subset of UDN patients with recessive disorders who experience seizures. Seizures do not have a singular cause, but instead may be the eventual result of many different biological and pathophysiological pathways. The specific pathways leading to seizure onset per patient are also often elusive; as many as half of global epilepsy cases have an unknown cause.^13^ Though antiepileptic drugs can reduce seizure burden in 60% to 70% of patients, these treatments minimize symptoms while rarely addressing the root biological cause.^14^ The overall phenotypic heterogeneity of cases within the UDN and enrichment of patients with seizures within this cohort provides the opportunity to explore the differing potential mechanisms that may predispose individuals to experience seizures.

For each UDN patient in our pilot with an identified strong candidate or diagnostic gene listed, we define “genotypically-similar” individuals as those who are seemingly healthy and carry at least one rare variant in the same gene. We select cohorts of genotypically-similar patients for each UDN patient from the UK Biobank (UKB), a large control dataset that contains both genetic and phenotypic patient information. We then examine the significant phenotype enrichments within each cohort to draw potential connections to the implicated body systems and biological mechanisms underlying the UDN patients’ conditions, including seizures.

## 1. Methods

### 2.1 Phenotype and genotype information for UDN patients

Comprehensive and relevant phenotype and genotype data for all patients enrolled in the UDN are uploaded to the central data repository managed by the UDN Data Management and Coordinating Center and made available to all UDN researchers through the UDN Gateway. These data often include standardized phenotype terms from the Human Phenotype Ontology (HPO) and candidate genes and variants that had been or are currently under investigation by the clinical teams in charge of each case. The genetic variants shared in the UDN Gateway could have been identified on a clinical sequencing report provided by the Clinical Laboratory Improvement Amendments (CLIA)-certified UDN Sequencing Core, through prior analyses or sequencing as documented in a patient’s medical records, or through internal site-specific protocols and pipelines. Genetic variants are marked as “candidate”, “solved” or “rejected.”

### 2.2 Curating set of recessive genes in seizure patients

We first filtered the 2366 patients enrolled in the UDN as of June 2022 to the subset of 622 patients annotated with at least one codified HPO seizure term. Our set of seizure terms was reviewed with a neurologist and consisted of 339 terms that were descendants of the seizure phenotype (HP:0001250) in the HPO ontology.^15^ We further filtered this list of patients to those with genes and variants listed in the UDN Gateway that the clinical team had classified as either candidate or confirmed causal. Some diagnoses in sporadic cases (i.e., two unaffected parents and no other affected family members) are due to single *de novo* variants likely operating in a dominant manner. To consider only recessive disease etiologies, we further filtered our set of patients to those with a recessive combination of putatively or known causal variants in the same gene, including homozygous alternate variants or compound heterozygous variant pairs. Note that in some recessively manifesting cases, only one of the requisite two causal variants may have been identified and listed in the UDN Gateway; these cases are excluded in our analysis. These filtering steps resulted in a set of 130 genes with a recessive combination of causal variants implicated in UDN patients experiencing seizures. All corresponding patient and gene counts for each filtering step are depicted in Figure 1.

**Figure 1.**
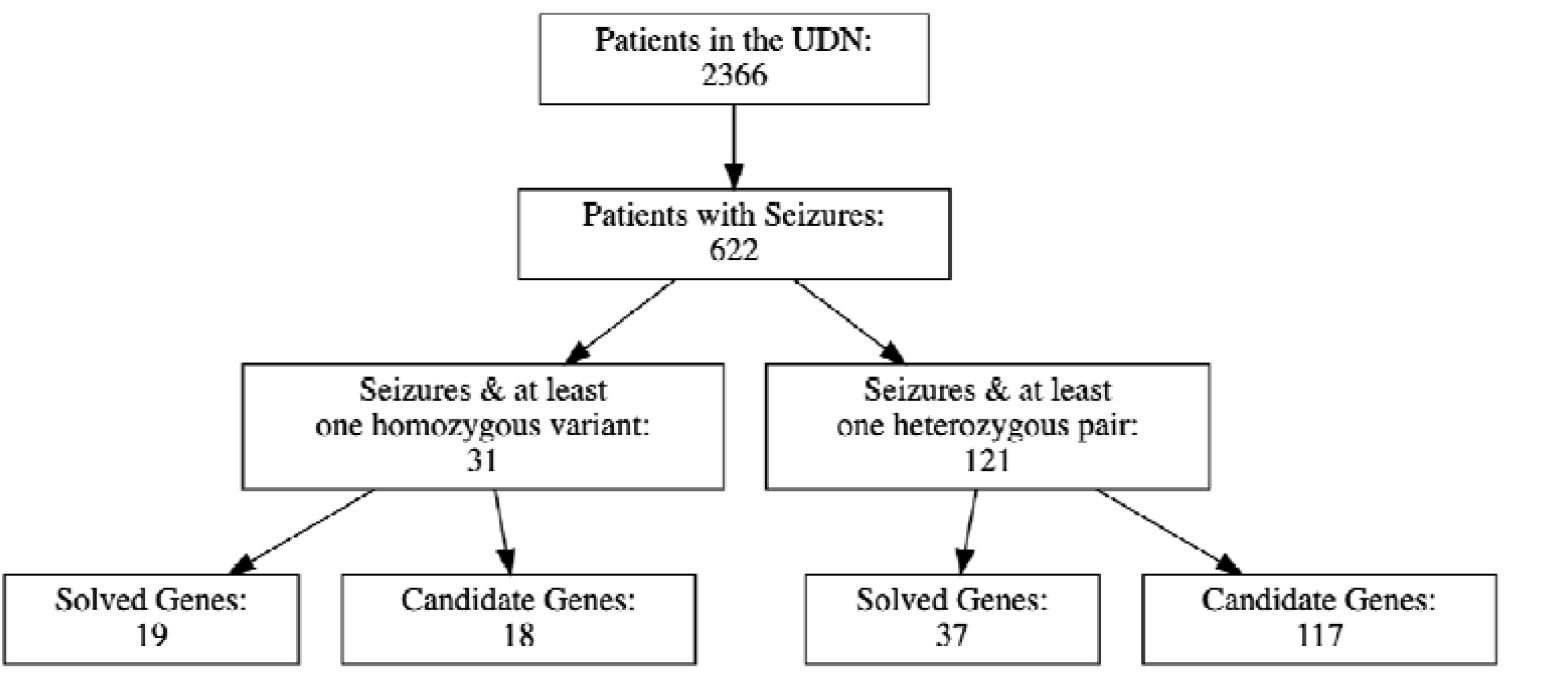
Selection process to identify genes of interest.

### 2.3 Identifying phenotypes enriched across genotypically-similar individuals

For each gene of interest in a UDN patient, we considered the corresponding subset of genotypically-similar individuals from the UKB who had whole exome sequencing and at least one rare variant in the same gene. We then queried Genebass, a resource that aggregates phenotypic information from 281,850 individuals with whole-exome sequencing in the UKB, to extract all phenotypes significantly associated with the aggregated rare variants per gene.^16^ Specifically, we considered phenotype association scores computed using SKAT-O, a method that first weights individual rare variants within a gene and then aggregates these variants per gene for better statistical power to compute phenotype associations using a burden test.^17^ We considered gene-based phenotype associations below the 2.5e-10 threshold to be significant.

### 2.4 Assessing significance between enriched phenotypes and UDN patient presentation

We compared the enriched phenotypes found in genotypically-similar UKB patients back to each starting UDN patient. We met with the clinicians and genetic counselors overseeing each patient’s case to discuss potential connections between the enriched phenotypes and potential biological mechanisms implicated in the UDN patients. The overall workflow is depicted in Figure 2.

**Figure 2.**
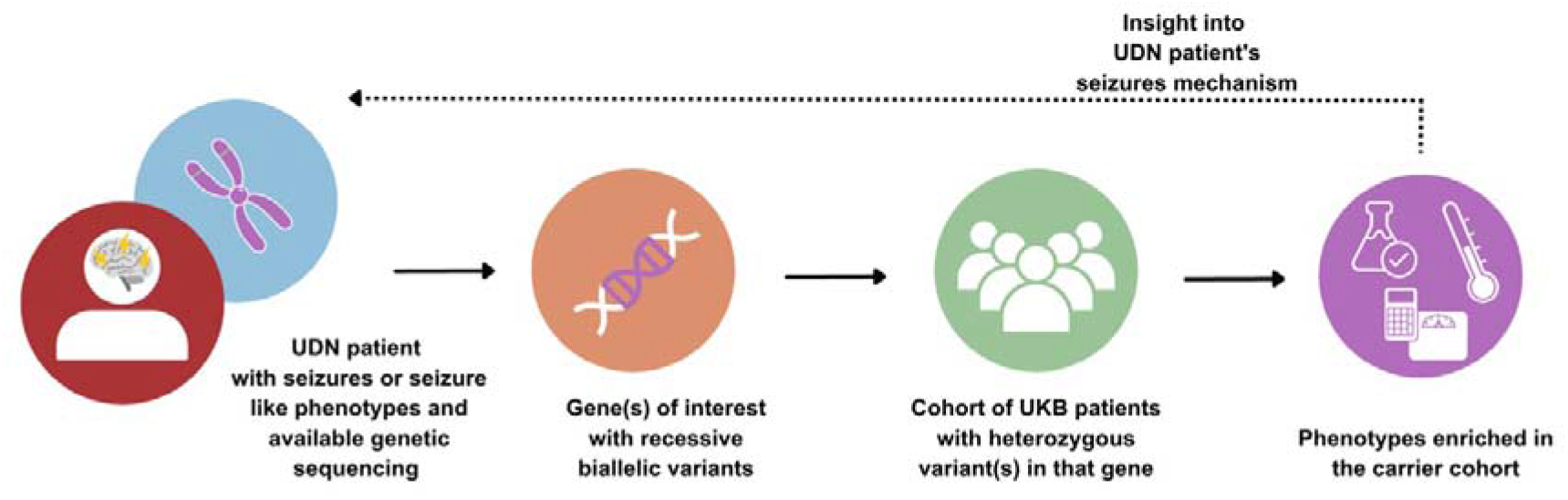
Workflow for deriving insight into rare disease patient’s seizure mechanism through cohort analysis.

## 2. Results

We first sought to identify genes where high impact variants may recessively manifest in rare disorders and seizures. To this end, we considered the subset of patients enrolled in the UDN who experienced seizures and had one or more listed genes harboring a recessive variant pair, occurring either as compound heterozygous variants or a homozygous variant (see Methods). This search resulted in a set of 238 unique variants falling into 130 genes across 109 patients. Variants are categorized by clinical teams into one of three causality statuses: candidate (n=159), solved (n=54), or rejected (n=25). Most variants are initially categorized as “candidate” and then recategorized as “solved” if experimental studies confirmed the variant’s causal impact on observed phenotype or additional patients with the same variant and compelling phenotypic overlap were identified through variant matchmaking services such as Matchmaker Exchange. Conversely, candidate variants are recategorized as “rejected” if functional studies demonstrated no phenotypic impact or additional individuals were found with the same variants but no overlapping symptoms. Computational tools can predict variant pathogenicity in the absence of experimental studies by considering protein structure, evolutionary conservation, and additional functionality measures (e.g., CADD, Polyphen, SIFT). As expected, we found that pathogenicity scores were significantly more deleterious for solved variants compared to candidate variants (p<0.001 across all three pathogenicity predictors, KS-test) and compared to rejected variants (p<0.001, KS-test, Figure 3). This suggests that the solved variants in our rare disease cohort likely correspond to strongly deleterious molecular impacts, and even carriers of one copy of these or similar variants may exhibit some disease-relevant symptoms.

**Figure. 3.**
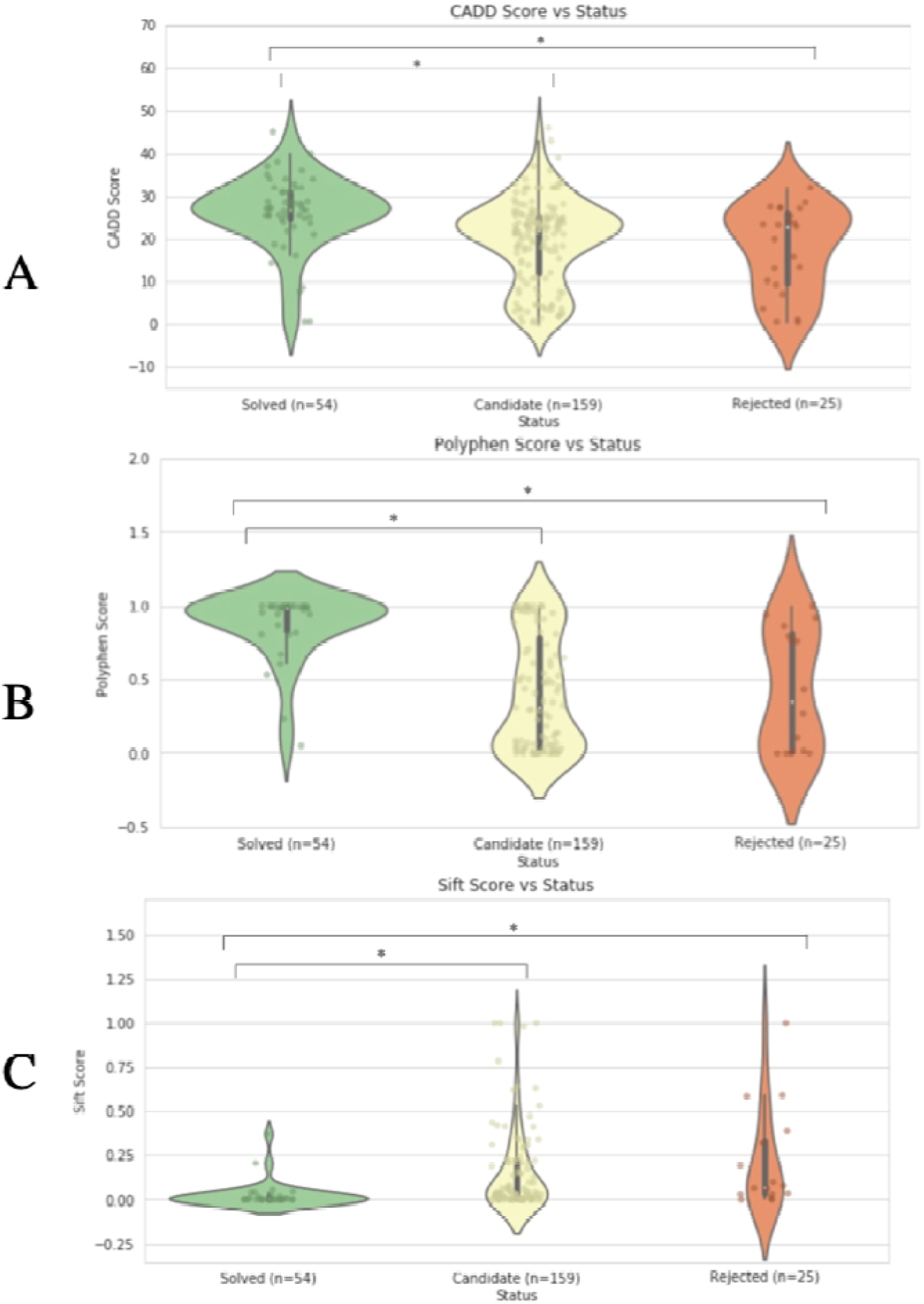
Deleteriousness vs Causality Status. A) CADD Score vs Status. B) Polyphen Score vs Status. C) Sift Score vs Status.

We next evaluated phenotypic enrichments across cohorts of individuals who are heterozygous for these specific variants or similar variants within each gene. For each of the 130 genes of interest in our UDN cohort, we considered individuals from the UKB with one or more variants in the same gene. We considered association values computed using SKAT-O per gene and provided through Genebass.^16^ Briefly, to increase statistical power to detect significant associations for rare variants that are harbored by only a few individuals each, SKAT-O assigns weights to rare variants based on their observed frequencies within the population and effect sizes, and then groups these variants within a gene to consider them together via a weighted burden test to compute phenotypic associations. Eight of the 130 genes tested had one or more phenotypes enriched below our significance threshold (Table 1). Two of these genes were removed from subsequent analyses upon further case investigation and discussions with the corresponding clinical teams. Specifically, DENND2C was determined to be unlikely to be causal by the UDN clinical team and the patient with variants in ATP7B was determined to not experience seizures.

**Table 1.**
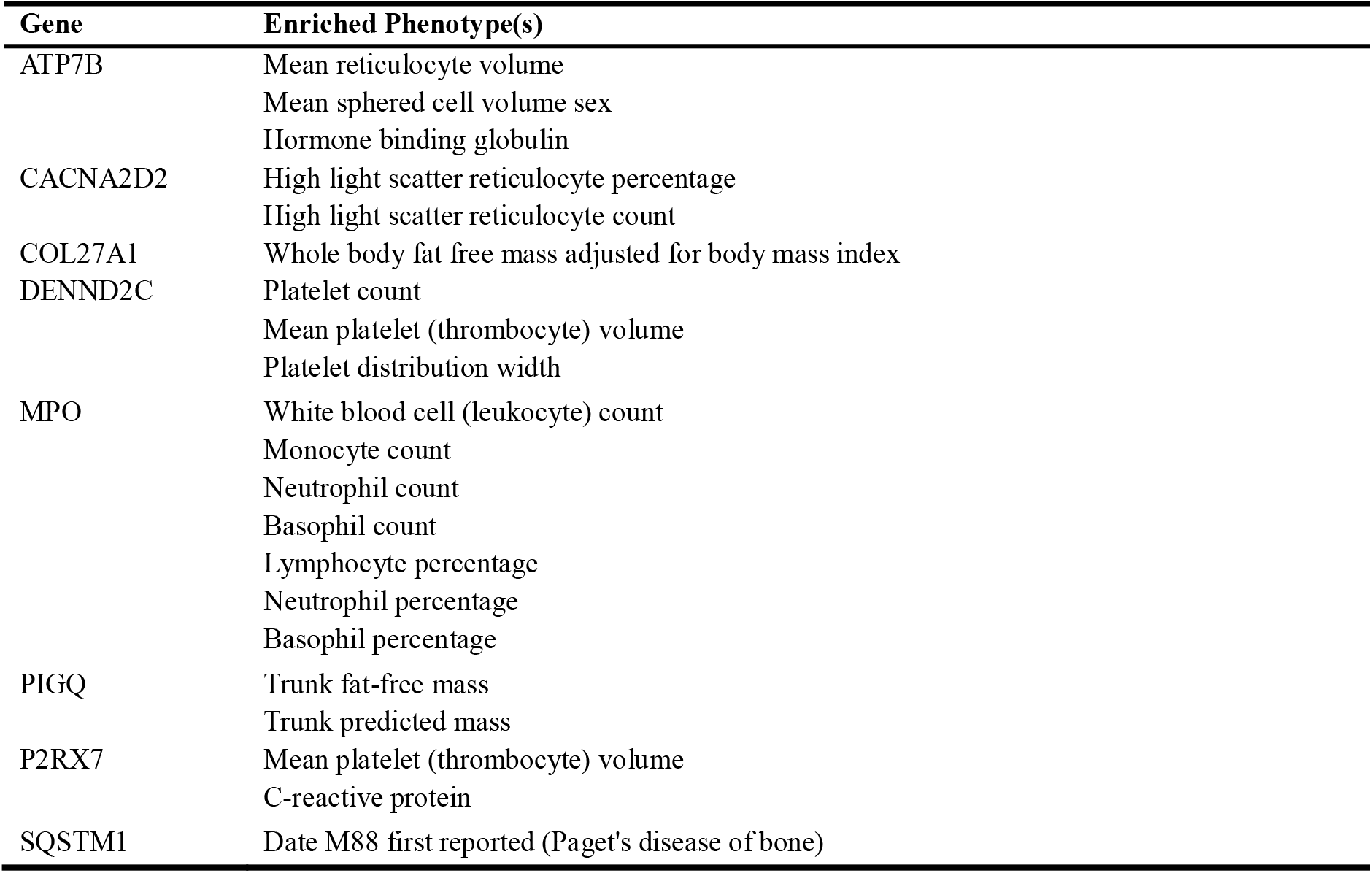
Enriched phenotypes for UK Biobank cohorts.

The phenotypes associated with the six remaining genes of interest—MPO, PIGQ, P2RX7, COL27A1, CACNA2D2, and SQSTM1—suggest plausible and potentially interesting mechanisms that could be relevant for each UDN patient’s condition. Intriguingly, these associated phenotypes primarily impacted the skeletal, circulatory, digestive, or immune systems rather than the nervous system (Figure 4). This result is in line with previous suggestions that seizures as a symptom may be an eventual and extreme result of a wide range of complex and heterogenous underlying biological causes that are not necessarily primarily neurological in nature. Indeed, the phenotypes associated with these candidate genes were not immediate and obvious precursors to seizures, but instead suggested multi-step pathways which could lead to seizures in our rare disease cases. Below we present six vignettes discussing phenotypes associated with each candidate gene in genotypically-similar cohorts from the UKB and how these phenotypes are related to the initial rare disease patient case.

**Figure 4.**
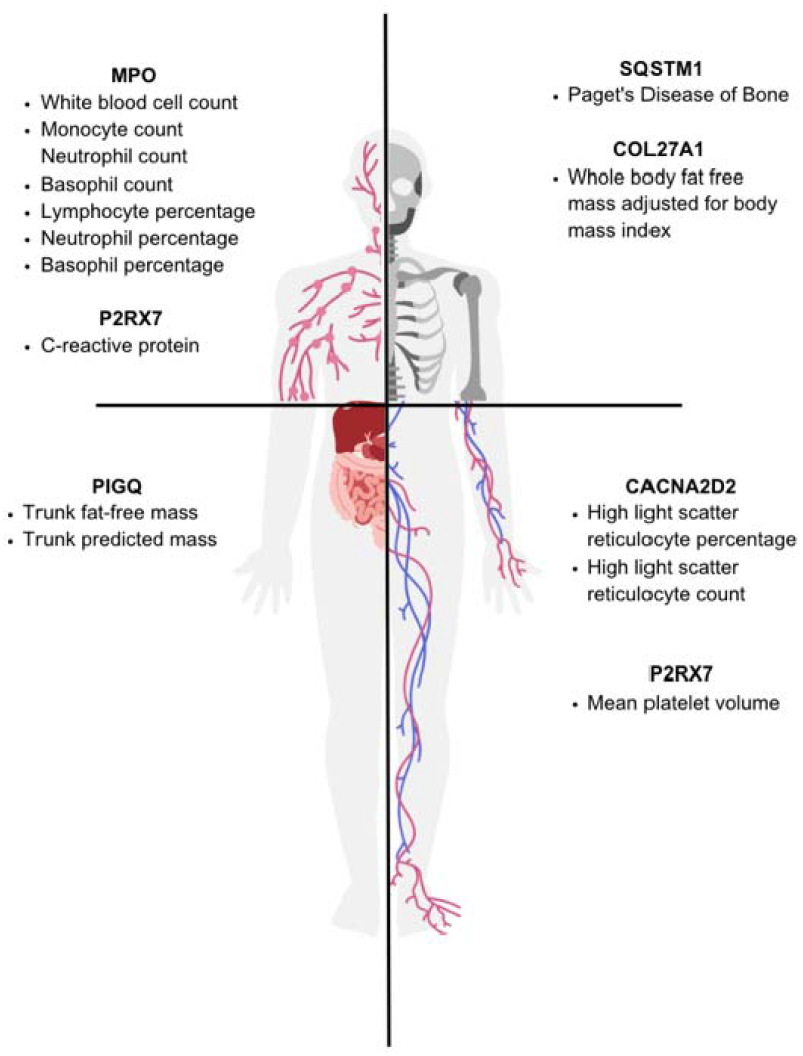
Enriched phenotypes and associated body systems involved.

### MPO

The phenotypes significantly associated with UKB participants with rare variants in the MPO gene were abnormal white blood cell (leukocyte) count, monocyte count, neutrophil count, basophil count, lymphocyte percentage, neutrophil percentage, and basophil percentage: all classic indicators of infection. The UDN patient with compound heterozygous variants in the MPO gene is highly sensitive to recurrent infections—including yeast infections and recurrent vaginal infections. These frequent infections are often accompanied by intense painful spells that result in insomnia. The clinical team assigned to this patient believes that this patient’s seizures are triggered by her insomnia.

Sleep deprivation is thought to promote seizure-causing interictal epileptiform discharges, or abnormal electrical activities that occur when a person is not actively experiencing a seizure.^18^ It has also been shown that increasing the duration a person sleeps at night decreases the likelihood that they will experience a subsequent seizure.^19^ As such, a connection between the patient’s insomnia and seizures is reasonable and suggests the patient’s seizures might be at least partially addressed by treating either the pain or insomnia. Though insomnia and seizures are only a part of the difficult and complicated conditions from which this patient suffers, her case is illustrative of how seizures can be consequence of an indirect pathway involving non-nervous body systems. Individuals who are genetically predisposed to abnormal infection response due to rare variants in the MPO gene may benefit from stricter or preemptive management of pain and sleep loss.

### P2RX7

An adolescent patient in the UDN suffers from bilateral tonic-clonic seizures and harbors a homozygous recessive variant in the P2RX7 gene. The phenotypes enriched among individuals with rare variants in P2RX7 include abnormal C reactive protein level and mean platelet volume. C reactive protein regulates platelet activation as part of the body’s inflammation response, and the rate of platelet production dictates mean platelet volume. Consequently, changes in these phenotypes are indictive of inflammation.^20,21^ Increased levels of C reactive protein have been found to be associated with epilepsy, suggesting a connection between inflammation and seizures.^22–24^ It is, however, difficult to use retrospective data to determine whether the inflammation is contributing to the seizures or is a result of recurrent seizures.

Both C reactive protein and mean platelet volume have also been associated with increased risk of thrombosis, or blood clots that obstruct normal blood flow.^20,21^ Abnormally increased platelet production can lead to blood clots and an increased risk for stroke.^25^ Patients who experience strokes are at heightened risk for seizures, as injuries to the brain can result in scar tissue and disruption to the brain’s electrical activity.^26^ Although dysregulation of platelet volume or C reactive protein levels may be related to seizures in general, there was limited evidence demonstrating abnormal platelet or C reactive protein levels in this particular UDN patient. However, these levels may have entered a normal range as a result of drug treatments for their symptoms.

### PIGQ

UKB participants with variants in the PIGQ gene were enriched for abnormal “trunk fat-free mass” and “trunk predicted mass”, two phenotype terms that are rarely used outside of the UKB. Trunk fat-free mass indicates abnormality in non-fat tissues such as organs in the abdominal region. Trunk predicted mass refers to general abnormalities in mass in the abdomen. The UDN rare disease patient with compound heterozygous variants in PIGQ has symptoms consistent with what could be detected as abnormal mass in abdominal organs—including chronic constipation, bulging pouches in the intestines, volvulus and bowel obstruction, and abdominal distention. Intriguingly, similar digestive symptoms as well as epilepsy have been found in a cohort of patients with variants in PIGQ. ^27^

Although PIGQ is seizure-associated, the biological mechanism underlying this connection, including any link between digestive and neurological symptoms, has not been well defined.^28^ PIGQ codes for Phosphatidylinositol Glycan Anchor Biosynthesis Class Q, a protein that helps synthesize glycosylphosphatidylinositol (GPI) anchors. GPI anchors attach proteins to the outer surface of cell membranes and are generally involved in cell-cell adhesion and the transport of molecules through cell membranes. ^29^ Though mutations in genes coding for GPI anchors have largely been associated with neurodevelopmental conditions, nervous system abnormalities are likely to impact the digestive system as well via the gut-brain axis. The gut-brain axis tightly links the nervous and digestive systems via neural, hormonal and immunological pathways to enable constant bidirectional communication between the gut and brain.^30^ Gastrointestinal disorders have been found to occur at higher rates in patients with epilepsy in general, and constipation is the second most common comorbidity in children with epilepsy.^31,32^ Indeed, chronic constipation results in a buildup of toxins that may be seizure-triggering.^32^ Variants in PIGQ could therefore contribute to epilepsy and indirectly to gastrointestinal abnormalities through a multi-step pathway.

### SQSTM1

UKB participants with variants in SQSTM1 were enriched for “Paget’s Disease of Bone”, a disorder characterized by joint stiffness, weakness, and abnormal bone growth resulting from new bone failing to effectively replace old bone.^33^ SQSTM1 has been identified as a potentially seizure-related gene as well, as abnormal skull growth can cause cerebellar compression and increased intercranial pressure that results in seizures in the extreme case.^28,34^ The UDN patient with compound heterozygous variants impacting SQSTM1 experiences abnormal movements and stiffness. Additionally, the patient’s skull circumference is smaller than the third percentile and had been characterized as infantile-onset microcephaly during the first year of life. As a result, the patient’s skull may have been even more compressed, putting them at greater risk for seizure activity.

### COL27A1

Individuals with variants in the COL27A1 gene were enriched for abnormal “whole body fat free mass adjusted for body mass index,” a UKB-specific phenotype term referring to abnormal bone or organ mass relative to body size. Consistently, the COL27A1 gene is associated with bone malformations, including hip dislocations, cleft pallet, scoliosis, and craniofacial differences.^35,36^ The UDN patient with compound heterozygous variants in COL27A1 has presented with each of these phenotypes. To address the patient’s craniosynostosis, or the premature fusing of some cranial sutures in the infant’s skull, surgery was required. Surgery to implant a shunt was then required to address the patient’s subsequent hydrocephalus, or the accumulation of excess fluid within the brain cavities. The connection between shunted hydrocephalus patients and seizures is well documented, with 20% to 50% of children with shunts also developing epilepsy.^37,38^ Shunt infections, one of the UDN patient’s symptoms, have been found to double the risk of developing epilepsy.^39^ The clinical team overseeing this case also feels that the patient’s epilepsy is likely a symptom resulting from the impacts on brain development. Overall, the compound heterozygous variants in COL27A1 are plausibly linked to the early-onset skeletal and skull abnormalities experienced by the UDN patient. These skeletal abnormalities may have led to seizure onset through the multi-step pathway described above, suggesting another biological mechanism potentially underlying seizures.

### CACNA2D2

The cohort of carriers for rare variants in CACNA2D2 were significantly associated with abnormal “high light scatter reticulocyte percentage” and “high light scatter reticulocyte count.” These two measurements are an indicator of a greater proportion of immature red blood cells than would be normally expected. Several of the UDN patient’s symptoms—looking spacy, seizures with cyanosis (bluish or purplish discoloration of the skin and mucous membranes), pallor (abnormally pale or whitish skin appearance), dark circles under the eyes, and lethargy—are all consistent with a lack of mature red blood cells.^40^ Complete blood count information that may have indicated abnormalities in red blood cell production were unavailable for this patient.

Although CACNA2D2 has independently been identified as a potentially seizure-related gene, any mechanism by which it is causatively related to seizures is unknown.^28^ Potential seizure-causing mechanisms related to a dysregulation in red blood cell production include oxygen deprivation to the brain, reduction of inhibitory neurotransmitters, and changes in the metabolism of neurons. ^41^

## 3. Discussion

Despite considerable scientific efforts, rare diseases remain exceedingly difficult to diagnose and understand mechanistically. Moreover, they are seldom treated effectively, and only ~6% of rare diseases have a treatment available at all.^42^ Interpreting findings in ultrarare disease patients whose conditions are not represented in published literature and who are too unique for traditional case-control statistical analyses requires unique approaches. Developing additional angles by which to assess these cases will expedite and enhance the diagnoses, understanding, and treatment of rare diseases. The availability of resources like the UK Biobank provides aggregated genotype and phenotype data at scale. Phenotypes observed across cohorts of “genotypically similar” individuals from these population-scale cohorts provide context to better understand small sets of unique patients and can illuminate new directions for future exploration, especially in complex cases where progress has otherwise stalled.

Over a quarter of all UDN patients have experienced seizures, underscoring the heterogeneity of mechanisms that may lead to seizure onset. In our pilot study presented here, we found phenotypic associations in UKB participants across four different body systems. These phenotypes suggested multistep pathways which might reasonably culminate in the occurrence of seizures. To eventually find disease modifying therapies, these understandings of the upstream seizure-causing processes are crucial.

Even when the disease-causing mechanisms in rare disease patients are understood, the development of treatment options for these “N-of-1” patients is often financially infeasible given the limited number of patients who may benefit. However, the same disease mechanisms implicated in ultrarare disorders may lead to similar, treatable symptoms in the general population, incentivizing the development of improved therapeutics. For example, individuals in the broader population who harbor rare variants in disease-causing genes may experience milder or later onset symptoms related to the severe symptoms that manifest in rare disease cases. Finding these links can inform positive interventions for both rare disease patients and for heterozygous variant carriers.

Given the unique and complex nature of each UDN case, further work is still necessary to conclusively validate any of these hypothesized pathways. In the six vignettes we presented here, the clinical teams overseeing each case accepted that the multi-step pathways leading to seizure onset were plausible, but there would be no easy way to confirm them or provide further evidence in support of or against the associations. The benefits of any additional testing, such as blood work or research assays, must be carefully considered given the high burden of medical tests and interventions these patients already undergo. Another important consideration is that many patients in the UDN overall and a subset of those included in our analyses here had multiple strong candidate diagnostic genes listed in the UDN Gateway. Although many disorders may be monogenic, some patients may have multiple genes implicated that together give rise to their set of manifesting symptoms. Moreover, not all genes are equally strong candidates in any given case. We discarded some genes from our analysis once we discovered through a meeting with the clinical team that a UDN patient had a different gene that was more likely to causative or relevant to the patient’s seizures.

The aims of this pilot study were two-fold: to investigate the etiology of seizures, but also to inspire future studies to interpret rare disease cases by leveraging large-scale biobank data. There are many directions to extend the pilot project presented here. First, it would be worthwhile to repeat a similar approach with all UDN patients or a subset of patients with a different initial phenotype with diverse, complex, and unknown causes. Second, this study could be further refined by considering and differentiating by variant functionality. In our work, UKB participants were considered genotypically similar if they harbored any rare variant in a gene of interest. However, variants at different positions in the same gene can have varying or even contradictory impact, so defining genotypically similar individuals as only those harboring variants with functionally similar impacts as the pathogenic variants in a rare disease patient is reasonable. However, restricting the cohort of “genotypically similar” individuals available for analysis this way is also likely to reduce statistical power for detecting significant associations. Third, incorporating candidate noncoding variants detectable from whole genome sequencing, rather than exome-only sequencing, would enable the consideration of many more variants and possible phenotypic associations. Although UDN patients have whole genome sequencing available, the UKB participants with precomputed phenotype associations available in Genebass only had exome data available.

A plethora of prior work has leveraged large-scale biobanks such as the UK Biobank to further our collective understanding of rare variants and their role in human disease. For instance, Patrick et al. identified patients within the UK Biobank diagnosed with rare diseases to identify new, potentially pathogenic variants for these diseases.^43^ Wang et al. and Jurgens et al. introduced foundational approaches associating rare variants to common diseases and phenotypes.^44,45^ In our work, we begin with patients already diagnosed with ultrarare or unique genetic conditions through N-of-1 analyses, and then leverage biobank data to further understand their conditions in an effort to shorten the therapeutic odyssey that follows the diagnostic odyssey for many of these patients.

The UDN provides both unique challenges and exciting opportunities for study. UDN patients are truly one-of-a kind, making their conditions difficult to understand and contextualize. Studying these complex conditions also highlights gaps in our overall biological understanding and pushes at the boundaries of current medical expertise. Analyzing these rare disease patients in the context of a larger cohort of genotypically similar biobank patients is one way to address the challenge of the conditions’ uniqueness and seizes the opportunity to better understand the underlying disease-causing mechanisms.

## Ethics and Inclusion Statement

The authors declare no competing interests. This work was performed in accordance with all ethical guidelines outlined in the NIH IRB #15HG0130. The study proposal and manuscript were approved by the Undiagnosed Diseases Network (UDN) Publications and Research Committee.

## Data Availability Statement

Deidentified genome data and corresponding phenotype data in the form of Human Phenotype Ontology (HPO) terms are regularly deposited in dbGaP (accession phs001232.v5.p2). Variant-level data, clinical significance and supporting evidence, demographic information, and phenotype information for all known pathogenic and confirmed diagnostic variants are regularly submitted to ClinVar. Identifiable patient data is under controlled access to protect patient privacy.

## Acknowledgments

We would like to thank the UDN, all of the participants in the UDN, as well as the clinical teams— including Brian Corner, Dr. John Phillips, Rebecca Spillmann, Dr. David Sweetser, Lauren Briere, Dr. James Orengo, and Jill Mokry—for their valuable insights. Research reported in this manuscript was supported by the National Institutes of Health (NIH) Common Fund grant U01HG007530 and the NIH National Institute of Neurological Disorders and Stroke (NINDS) grants U2CNS132415 and R00NS114850. The content is solely the authors’ responsibility and does not necessarily represent the official views of the National Institutes of Health.

---

**Supplementary Note 1.** Members of the Undiagnosed Diseases Network

Maria T. Acosta, David R. Adams, Ben Afzali, Ali Al-Beshri, Eric Allenspach, Aimee Allworth, Raquel L. Alvarez, Justin Alvey, Ashley Andrews, Euan A. Ashley, Carlos A. Bacino, Guney Bademci, Ashok Balasubramanyam, Dustin Baldridge, Erin Baldwin, Jim Bale, Michael Bamshad, Deborah Barbouth, Pinar Bayrak-Toydemir, Anita Beck, Alan H. Beggs, Edward Behrens, Gill Bejerano, Hugo J. Bellen, Jimmy Bennett, Jonathan A. Bernstein, Gerard T. Berry, Anna Bican, Stephanie Bivona, Elizabeth Blue, John Bohnsack, Devon Bonner, Nicholas Borja, Lorenzo Botto, Lauren C. Briere, Elizabeth A. Burke, Lindsay C. Burrage, Manish J. Butte, Peter Byers, William E. Byrd, Kaitlin Callaway, John Carey, George Carvalho, Thomas Cassini, Sirisak Chanprasert, Hsiao-Tuan Chao, Ivan Chinn, Gary D. Clark, Terra R. Coakley, Laurel A. Cobban, Joy D. Cogan, Matthew Coggins, F. Sessions Cole, Brian Corner, Rosario I. Corona, William J. Craigen, Andrew B. Crouse, Vishnu Cuddapah, Precilla D’Souza, Hongzheng Dai, Kahlen Darr, Surendra Dasari, Joie Davis, Margaret Delgado, Esteban C. Dell’Angelica, Katrina Dipple, Daniel Doherty, Naghmeh Dorrani, Jessica Douglas, Emilie D. Douine, Dawn Earl, Lisa T. Emrick, Christine M. Eng, Cecilia Esteves, Kimberly Ezell, Elizabeth L. Fieg, Paul G. Fisher, Brent L. Fogel, Jiayu Fu, William A. Gahl, Rebecca Ganetzky, Emily Glanton, Ian Glass, Page C. Goddard, Joanna M. Gonzalez, Andrea Gropman, Meghan C. Halley, Rizwan Hamid, Neal Hanchard, Kelly Hassey, Nichole Hayes, Frances High, Anne Hing, Fuki M. Hisama, Ingrid A. Holm, Jason Hom, Martha Horike-Pyne, Alden Huang, Yan Huang, Anna Hurst, Wendy Introne, Gail P. Jarvik, Suman Jayadev, Orpa Jean-Marie, Vaidehi Jobanputra, Oguz Kanca, Yigit Karasozen, Shamika Ketkar, Dana Kiley, Gonench Kilich, Eric Klee, Shilpa N. Kobren, Isaac S. Kohane, Jennefer N. Kohler, Bruce Korf, Susan Korrick, Deborah Krakow, Elijah Kravets, Seema R. Lalani, Christina Lam, Brendan C. Lanpher, Ian R. Lanza, Kumarie Latchman, Kimberly LeBlanc, Brendan H. Lee, Kathleen A. Leppig, Richard A. Lewis, Pengfei Liu, Nicola Longo, Joseph Loscalzo, Richard L. Maas, Ellen F. Macnamara, Calum A. MacRae, Valerie V. Maduro, AudreyStephannie Maghiro, Rachel Mahoney, May Christine V. Malicdan, Rong Mao, Ronit Marom, Gabor Marth, Beth A. Martin, Martin G. Martin, Julian A. Martínez-Agosto, Shruti Marwaha, Allyn McConkie-Rosell, Ashley McMinn, Matthew Might, Mohamad Mikati, Danny Miller, Ghayda Mirzaa, Breanna Mitchell, Paolo Moretti, Marie Morimoto, John J. Mulvihill, Lindsay Mulvihill, Mariko Nakano-Okuno, Stanley F. Nelson, Serena Neumann, Dargie Nitsuh, Donna Novacic, Devin Oglesbee, James P. Orengo, Laura Pace, Stephen Pak, J. Carl Pallais, Neil H. Parker, LéShon Peart, Leoyklang Petcharet, John A. Phillips III, Filippo Pinto e Vairo, Jennifer E. Posey, Lorraine Potocki, Barbara N. Pusey Swerdzewski, Aaron Quinlan, Daniel J. Rader, Ramakrishnan Rajagopalan, Deepak A. Rao, Anna Raper, Wendy Raskind, Adriana Rebelo, Chloe M. Reuter, Lynette Rives, Lance H. Rodan, Martin Rodriguez, Jill A. Rosenfeld, Elizabeth Rosenthal, Francis Rossignol, Maura Ruzhnikov, Marla Sabaii, Jacinda B. Sampson, Timothy Schedl, Lisa Schimmenti, Kelly Schoch, Daryl A. Scott, Elaine Seto, Vandana Shashi, Emily Shelkowitz, Sam Sheppeard, Jimann Shin, Edwin K. Silverman, Giorgio Sirugo, Kathy Sisco, Tammi Skelton, Cara Skraban, Carson A. Smith, Kevin S. Smith, Lilianna Solnica-Krezel, Ben Solomon, Rebecca C. Spillmann, Andrew Stergachis, Joan M. Stoler, Kathleen Sullivan, Shamil R. Sunyaev, Shirley Sutton, David A. Sweetser, Virginia Sybert, Holly K. Tabor, Queenie Tan, Arjun Tarakad, Herman Taylor, Mustafa Tekin, Willa Thorson, Cynthia J. Tifft, Camilo Toro, Alyssa A. Tran, Rachel A. Ungar, Adeline Vanderver, Matt Velinder, Dave Viskochil, Tiphanie P. Vogel, Colleen E. Wahl, Melissa Walker, Nicole M. Walley, Jennifer Wambach, Michael F. Wangler, Patricia A. Ward, Daniel Wegner, Monika Weisz Hubshman, Mark Wener, Tara Wenger, Monte Westerfield, Matthew T. Wheeler, Jordan Whitlock, Lynne A. Wolfe, Heidi Wood, Kim Worley, Shinya Yamamoto, Zhe Zhang, Stephan Zuchner

